# Community factors and excess mortality in first wave of the COVID-19 pandemic

**DOI:** 10.1101/2020.11.19.20234849

**Authors:** Bethan Davies, Brandon L Parkes, James Bennett, Daniela Fecht, Marta Blangiardo, Majid Ezzati, Paul Elliott

## Abstract

Risk factors for increased risk of death from Coronavirus Disease 19 (COVID-19) have been identified^1,2^ but less is known on characteristics that make communities resilient or vulnerable to the mortality impacts of the pandemic. We applied a two-stage Bayesian spatial model to quantify inequalities in excess mortality at the community level during the first wave of the pandemic in England. We used geocoded data on all deaths in people aged 40 years and older during March-May 2020 compared with 2015-2019 in 6,791 local communities. Here we show that communities with an increased risk of excess mortality had a high density of care homes, and/or high proportion of residents on income support, living in overcrowded homes and/or high percent of people with a non-White ethnicity (including Black, Asian and other minority ethnic groups). Conversely, after accounting for other community characteristics, we found no association between population density or air pollution and excess mortality. Overall, the social and environmental variables accounted for around 15% of the variation in mortality at community level. Effective and timely public health and healthcare measures that target the communities at greatest risk are urgently needed if England and other industrialised countries are to avoid further widening of inequalities in mortality patterns during the second wave.

---

Excess mortality during the COVID-19 pandemic is the combination of deaths caused, or contributed to, by infection with SARS-CoV-2 plus deaths that resulted from the widespread behavioural, social and healthcare changes that accompanied national responses to the emergency^1,3-5^. England has experienced one of the highest death tolls from COVID-19 in the industrialised world, far beyond what would be expected from its underlying health status and factors like obesity^1,2,6,7^. COVID-19 may substantially widen existing national health inequalities as the direct and indirect impacts of the pandemic may disproportionately effect the population groups with the highest healthcare utilisation: the elderly, people with chronic health conditions, people from a minority ethnic background and people who live in more deprived areas^2,8^.

Rates of diagnosed SARS-CoV-2 infections and deaths among people with confirmed infection vary substantially across England^9^. But neither local variations in all-cause mortality associated with the pandemic, nor their community determinants, are well understood; population density and urbanisation are often mentioned as major contributory factors in urbanised industrialised countries^10,11^. Here, we analysed geocoded data on all-cause mortality at ages 40 years and over for 6,791 local communities (Middle Super Output Areas [MSOAs]; median population 7,956 in 2018, median area 3.04 km^2^, Extended Data Table 1) in England to quantify local variations in excess mortality in the first wave of the pandemic, from 1 March to 31 May 2020, and to identify the community characteristics associated with these patterns.

From 1 March to 31 May 2020, 171,294 people at ages 40 years and over died in England, compared with a mean of 121,358 deaths in the same period in 2105-2019, equivalent to 49,936 excess deaths. Compared with 2015-2019, a greater proportion of the deaths in 2020 were in men, in care homes and a smaller proportion occurred in hospitals (Fig. 1).

**Fig. 1.**
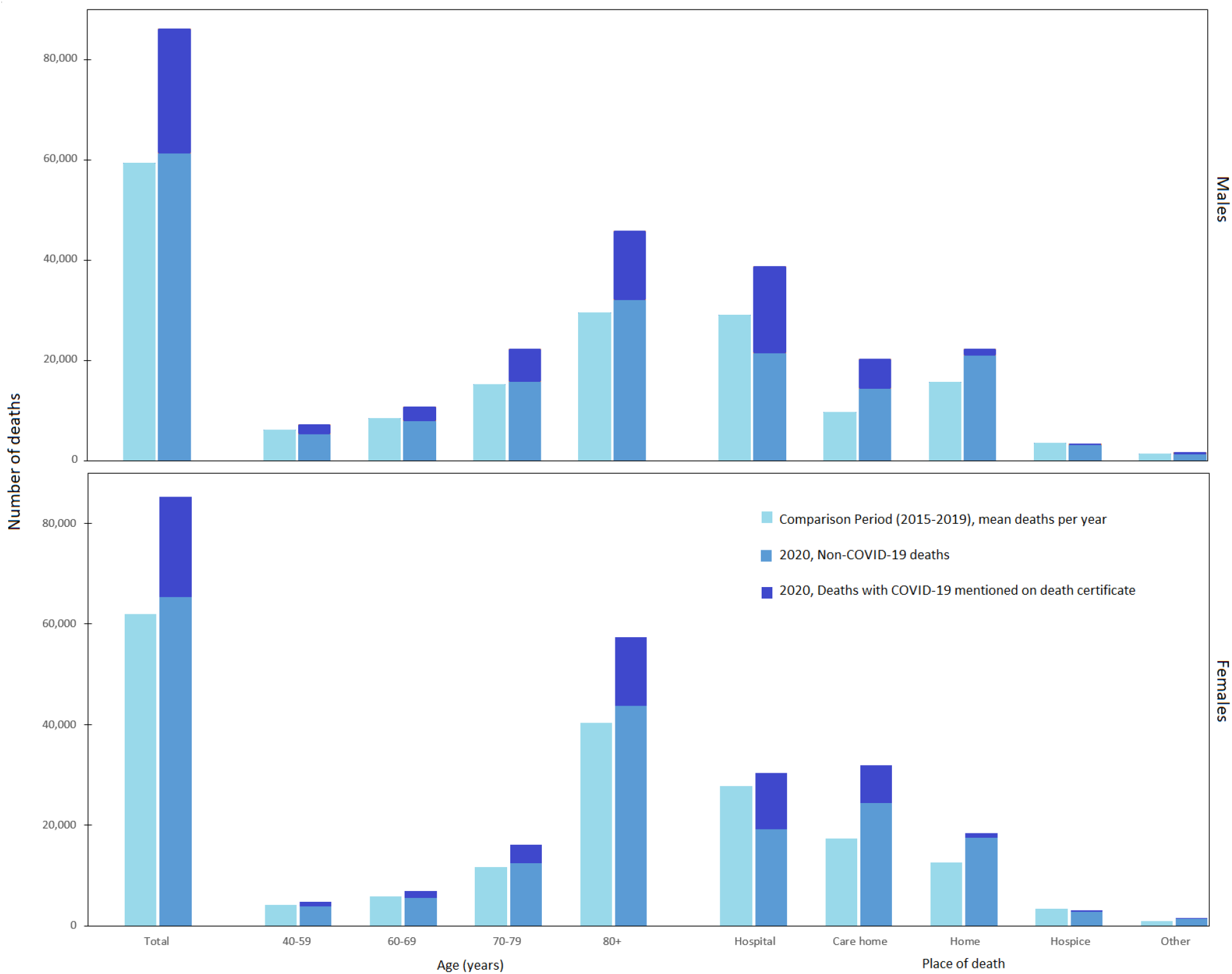
Distributions of deaths by sex, age and place of death. Study period: 1 March to 31 May 2020. Comparison period: 1 March to 31 May 2015-2019.

Because communities (MSOAs) are small, we used a Bayesian spatial model to obtain stable estimates of excess death rates based on data for each community and those of its neighbours to reduce uncertainty (<u>Methods</u>). The spatial model included terms for potential community determinants of mortality: percent population on income support as a marker of area poverty; population density; percent who are non-White; and percent population living in overcrowded homes. We also included air pollution, namely annual average nitrogen dioxide (NO_2_) and fine particulate matter (PM_2.5_) and number of care homes per 1,000 population. Data sources and definitions of these variables are detailed in <u>Methods</u>. Each variable was divided into quintiles of the distribution to allow for non-linear relationships (Extended Data Table 2).

All but 360 communities in men and 668 in women had higher mortality in 2020 than expected based on prior years (Fig. 2) with a posterior probability of increased mortality of at least 90% in 4,240 (62.4%) communities in men and 3,166 (46.7%) in women. Of these, mortality more than doubled in 697 (10.3%) and more than tripled in 18 (0.26%) communities in men and in 498 (7.3%) and 15 (0.22%) communities respectively in women (Extended Data Table 3).

**Fig. 2.**
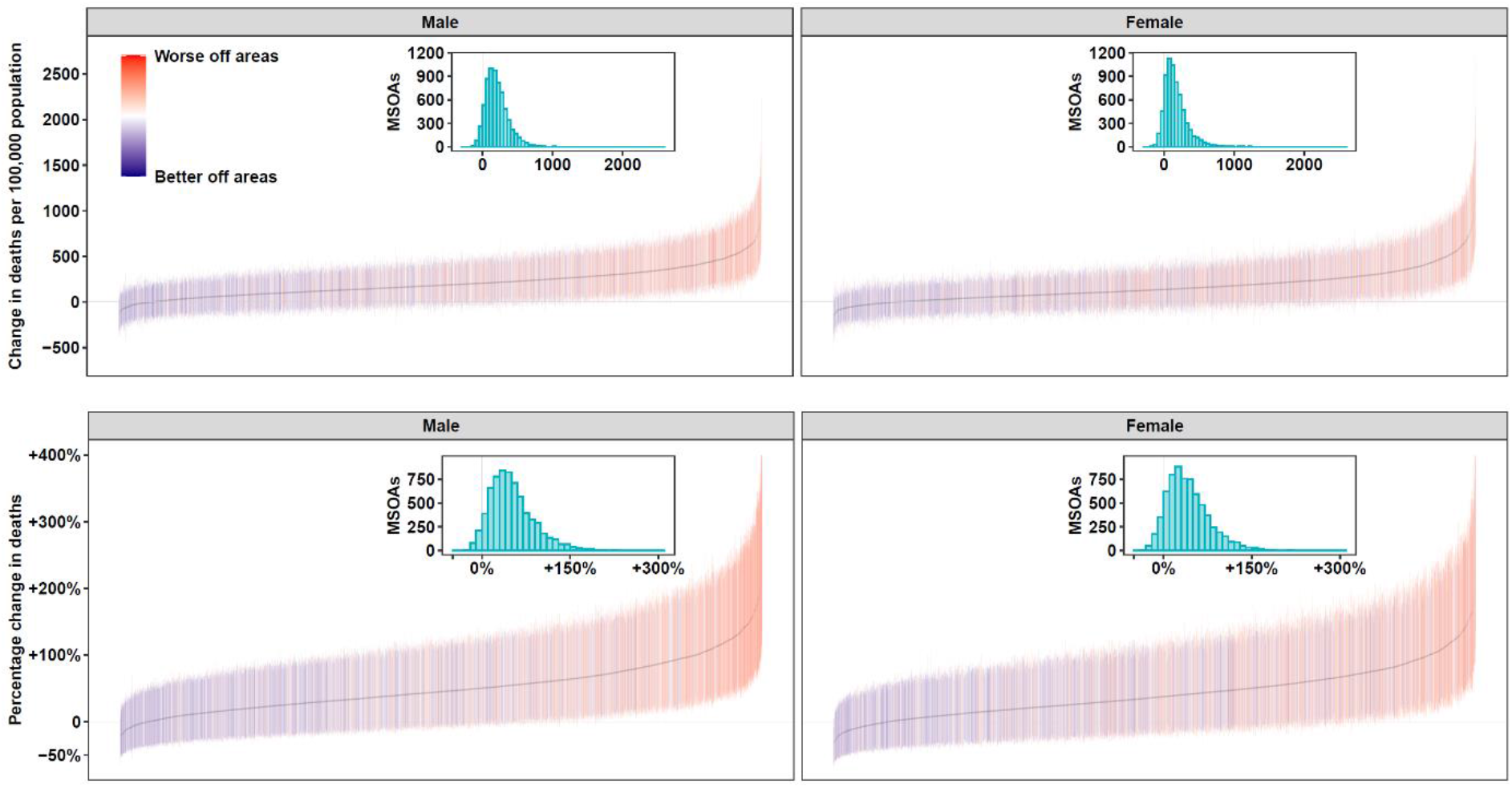
Excess mortality (with 95% credible intervals) from 1 March to 31 May 2020 compared to the same period for the preceding five years, for middle super output areas (MSOAs) in England. (A) Excess deaths per 100,000 people aged 40 years and over in 2020 compared to the average for the same period for the preceding five years. (B) Percent increase in death rates in 2020 compared to the average for the same period for the preceding five years. Colouring indicates the sum (by quintiles) of community characteristics of the MSOAs: % population on income support; population density; % population non-White; % population living in overcrowded homes; air pollution (NO_2_ and PM_2.5_); care homes per 1,000 population. Inserts are histograms of the distribution of (A) excess deaths per 100,000 people aged 40 years and over, and (B) percent increase in death rates across MSOAs.

The communities with an increase in mortality were spread across the country with the lowest increases in remote rural areas (Fig. 3). The largest increases in mortality were concentrated in London^12^, especially for men; for women high excess mortality also occurred in suburban areas. In men, 30.8% of variation in excess mortality was explained by local clustering but only 18.4% in women, suggesting greater correlation in excess mortality between neighbouring areas for men than women. On average, communities with large increases in mortality tended to have greater social and environmental deprivation than those with small increases (Fig. 2).

**Fig. 3.**
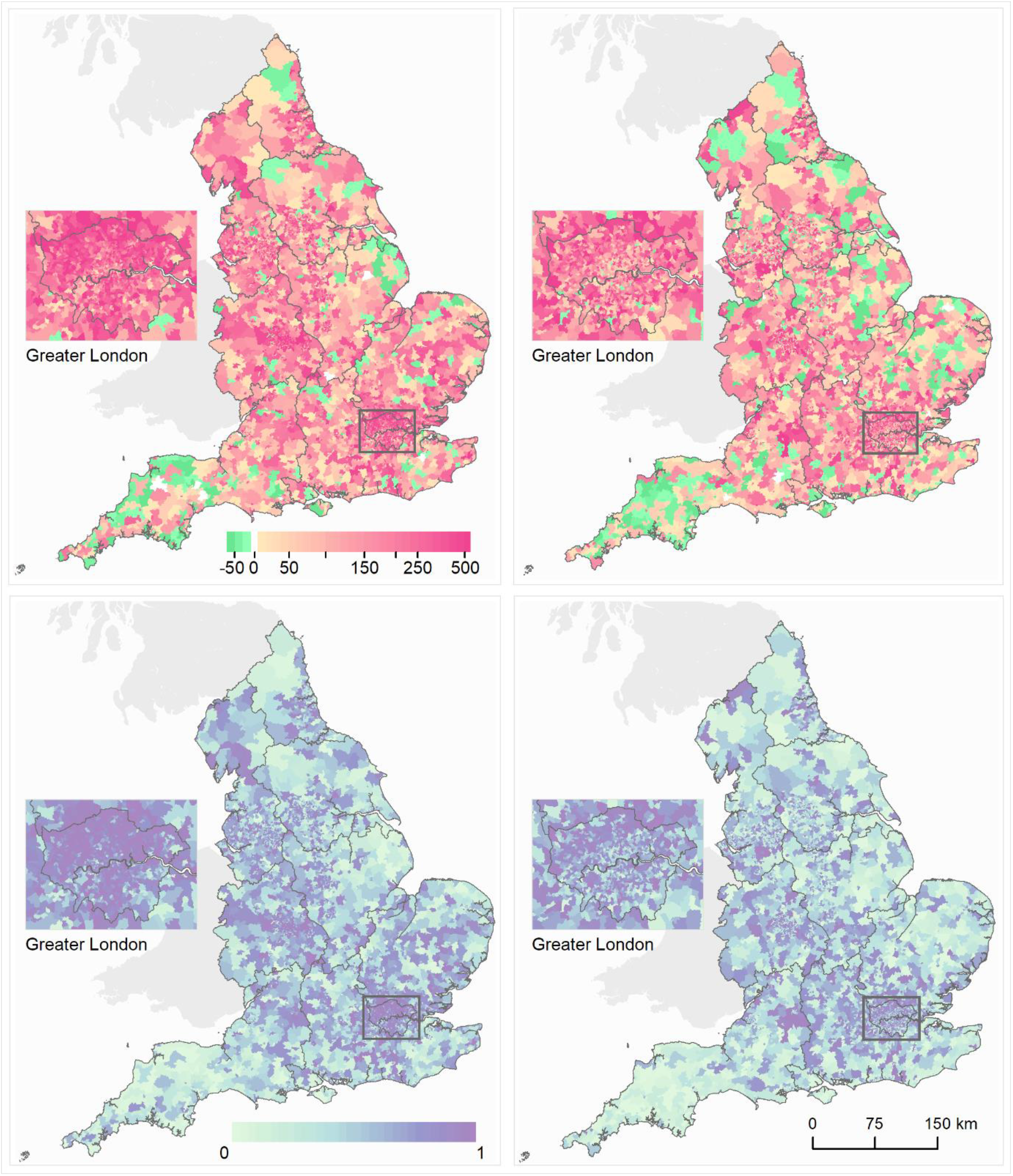
Maps of middle super output areas (MSOAs) in England showing excess deaths per 100,000 people aged 40 years and over. (A) Excess deaths per 100,000 males (left)/females (right) from 1 March to 31 May 2020 compared to the same period for the preceding five years. (B) Posterior probability that excess deaths > 0. Community characteristics of the MSOAs were: % population on income support; population density; % population non-White; % population living in overcrowded homes; air pollution (NO_2_ and PM_2.5_); care homes per 1,000 population. We map the posterior probability which measures the extent to which an estimate of excess/fewer deaths is likely to be a true increase/decrease. Where the entire posterior distribution of estimated excess deaths for an MSOA is greater than zero, there is a posterior probability of ∼1 of a true increase, and conversely where the entire posterior distribution is less than zero there is a posterior probability of ∼0 of a true increase. This posterior probability would be ∼0.5 in an MSOA in which an increase is statistically indistinguishable from a decrease (Extended Data Table 3).

The combination of a large relative increase in mortality and a high baseline (i.e. pre-pandemic) death rate meant that men in 2,240 communities and women in 1,544 communities experienced 250 or more excess deaths per 100,000 people aged 40 years and over compared with the prior years; in 333 communities for men and 305 for women, the excess mortality burden was at least 500 per 100,000 people. The large variation in excess death rates meant that 25% of all excess deaths during the pandemic occurred in only 9.4% of communities for men and 9.1% for women, and one half of excess deaths occurred in 24.3% and 24.2% of communities, respectively. Excess deaths per 100,000 people were only moderately correlated between men and women (Extended Data Fig. 1 and 2).

Each of the community characteristics considered was individually (i.e. in univariate analysis) associated with excess deaths during the pandemic in graded fashion across quintiles (Fig. 4, Extended Data Table 4). However, there were strong inter-correlations between some variables; for example, Kendall’s Tau was 0.67 and 0.55 between percent non-White population and levels of NO_2_ and PM_2.5_ respectively (Extended Data Table 5). When the community characteristics were considered jointly in multivariable analyses, air pollution and population density were no longer associated with excess deaths, contrary to reports in some studies that air pollution is a contributory factor for COVID-19 deaths (Extended Data Table 6)^13-16^. Relationships with income support, percent non-White population and overcrowded homes persisted, although were attenuated – with a ∼10% higher rate across quintiles for men, and somewhat weaker associations for women. The relationship with care home density, even after accounting for the other variables, remained strong, with a ∼22% higher excess death rate for men and ∼27% for women in communities with the highest compared to lowest density of care homes; many of these deaths were not assigned to COVID-19^17^. Overall, the community variables accounted for 17.6% of the variation in mortality at community level in men and 14.9% in women (Extended Data Table 6). Sensitivity analyses with different smoothing parameters, excluding deaths in care homes, and combining data for men and women, did not materially alter our findings (Extended Data Table 7 a-e)

**Fig. 4.**
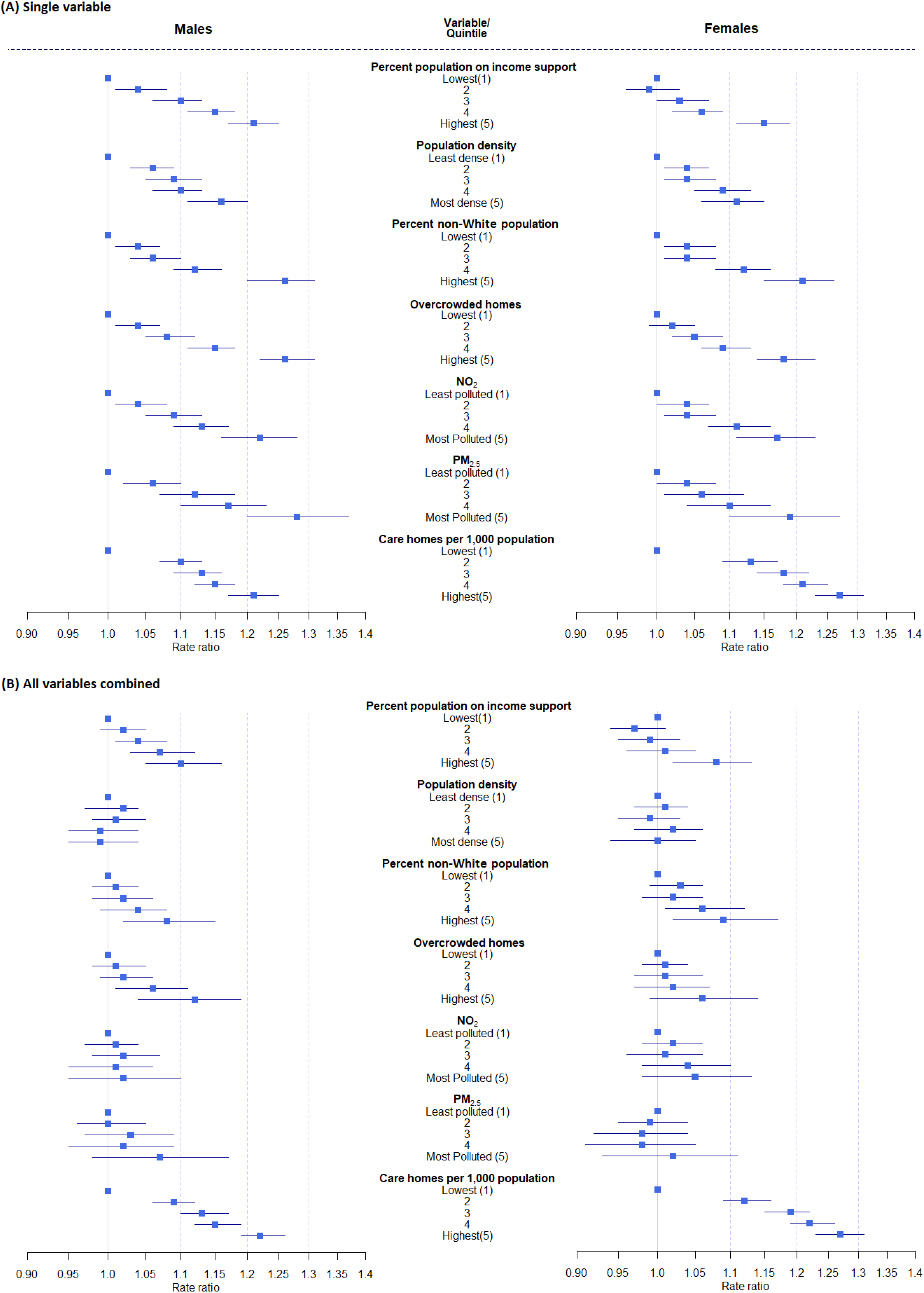
The relationship between community characteristics of middle super output areas (MSOAs) in England and excess mortality from 1 March to 31 May 2020 compared to the same period for the preceding five years. (A) Univariable relationship between each characteristic and excess mortality; (B) Multivariable relationship between characteristic and excess mortality after adjustment for the other characteristics. Proportional increase in death rates shown as rate ratios (with 95% credible intervals) for quintiles of the distributions relative to lowest quintile.

Our study has strengths and limitations. We included excess mortality from all causes, not just deaths coded to COVID-19. This should give the most complete picture of the effects of the pandemic on mortality and is comparable across geographies, as it is not dependent on availability of testing or diagnostic facilities nor variations in national or local coding practices. Not only could COVID-19 deaths have been wrongly ascribed to other causes but deaths from other causes may have been affected by the switching of healthcare resources to deal with the pandemic^6,8,18-20^. Although we accounted for population changes in the communities during the study period, population estimates at this scale were only available to 2018 and were extrapolated for the subsequent years. We used mortality data for the same three months of the year (March to May) over the previous five years to estimate the expected numbers of deaths in those months during 2020. But factors like temperature may modify the number of deaths observed. In addition, we used data from the last national census in 2011 to obtain information on sociodemographic characteristics of communities. To the extent that there have been demographic changes in the nine years since then, this may have led to misclassification of areas with respect to their community characteristics.

Our finding on the importance of care home density as a predictor of local excess mortality is consistent with the policy in the National Health Service to discharge up to 15,000 medically fit inpatients to avoid hospitals becoming overwhelmed^21^. It is likely that many of the elderly individuals discharged in this way will have needed support from social care services (including care homes) on discharge^22^ and may have not been tested for the SARS-CoV-2 virus prior to discharge^23^. Our study also underlines the associations between excess mortality and poverty, non-White ethnicity^24,25^ and overcrowded housing^26,27^at the community-level. Those living in poor communities and overcrowded homes have fewer opportunities for adopting measures that reduce transmission^28^, have higher exposure to infection at work or may be more restricted in terms of accessing healthcare for COVID-19 and other conditions^29^. Recent and ongoing research indicates that higher risks associated with ethnicity may at least in part reflect higher levels of overcrowding and poverty (adjusted for in our analysis), higher representation in frontline jobs in the health and care sector^12,30^, slower access to and utilisation of healthcare^26,29,31,32^, and possibly higher rates of co-morbidities such as diabetes and obesity^2,33,34^. Further research to understand the pathways underpinning these associations is needed to inform long-term strategy to tackle the social and environmental drivers of inequality that may have contributed to differential mortality during the pandemic.

In the short-term, as industrialised countries in Europe and elsewhere confront the pandemic’s second wave, in addition to more attention to protecting care home residents and workers^35^, the response from many governments has been either a national lockdown or a tiered lockdown applied primarily to cities^36^. England has now entered its second national lockdown. Lockdowns in the first wave were highly effective at driving down the rates of new infection, but they are not sustainable ^4,37-39^. Therefore, the immediate priority is to continue to strengthen public health systems to ensure they have the capacity, in real-time, to test and diagnose newly infected individuals; identify their contacts; provide self-isolation and quarantine advice; and undertake national surveillance to inform the evolving policy response^40^. In parallel, economic interventions that support job security and provide financial compensation to low-paid workers required to self-isolate are essential to support population-level compliance with public health advice^41,42^.

In summary, in one of the worst affected industrialised countries in the first wave of the COVID-19 pandemic, we found substantial community-level variation in excess mortality, ranging from small declines to tripling in mortality in some areas. Although at first glance the high increases are more evident in cities, population density itself does not appear to be a driver of excess mortality; rather excess mortality risks are related to poverty, overcrowded homes and non-White ethnicity, parallel to large impacts in communities where care homes are located. While we found that these community factors and geographical clustering contributed independently to patterns of excess mortality, a large proportion of the variance remained unexplained. This underlines the importance of using real-time surveillance to identify local outbreaks and target public health resource according to need. A robust public health response is essential if England and other industrialised countries are to control the transmission of SARS-CoV-2 and avoid further widening of inequalities in mortality patterns during the second wave.

## Supporting information

STROBE checklist

## Data Availability

Data availability:
No identifiable information will be shared with any other organisation. SAHSU does not have permission to supply data to third parties. Individual mortality data can be requested through the Office for National Statistics (https://www.ons.gov.uk/).
Mid-year population estimates can be downloaded from https://www.ons.gov.uk/peoplepopulationandcommunity/populationandmigration/populationestimates/datasets/middlesuperoutputareamidyearpopulationestimates.
English Index of Multiple Deprivation data can be downloaded from https://www.gov.uk/government/statistics/english-indices-of-deprivation-2019.
2011 Census data can be downloaded from https://www.ons.gov.uk/census/2011census/2011censusdata.
Modelled air pollution data (NO2 & PM2.5) can be downloaded from https://uk-air.defra.gov.uk/data/pcm-data.
Locations data of care homes can be downloaded from https://covid19.esriuk.com/datasets/e4ffa672880a4facaab717dea3cdc404_0.
Code availability:
The computer code written in R51 for the two stages of Bayesian models used in this work is available on request.

https://www.gov.uk/government/statistics/english-indices-of-deprivation-2019

https://www.ons.gov.uk/census/2011census/2011censusdata

https://uk-air.defra.gov.uk/data/pcm-data

https://covid19.esriuk.com/datasets/e4ffa672880a4facaab717dea3cdc404_0

## Methods

### Data sources

The study uses data held by the UK Small Area Health Statistics Unit (SAHSU), obtained from the Office for National Statistics (ONS). The ONS individual mortality data included date of death, date of registration of death, place of residence of the deceased, place of death (e.g. hospital, hospice, care home, at home), and International Classification of Diseases tenth revision (ICD10) codes of the underlying cause of death. Annual population was from ONS mid-year population estimates by age and sex for communities (MSOAs) in England, 2015 to 2018. 2019 MSOA populations were estimated by distributing 2019 local authority district populations according to MSOA shares in 2018. No 2020 population data are yet available and 2019 estimates were used instead.

### Characteristics of MSOAs (communities)

To investigate the association of community characteristics with excess mortality we included the following data at MSOA level:

- Income deprivation: Proportion of the population (adults and children, including asylum seekers) on government assistance due to low income and unemployment^43^.
- Population density: Number of people per square kilometre from 2019 mid-year population estimates, as described above.
- Ethnicity: Percentage of the population of ethnic origin other than White from 2011 census data^44^.
- Housing: Percentage of overcrowded households defined as households with at least one fewer bedroom as required based on the number of household members and their relationship to each other, from 2011 census data^44^.
- Air pollution: Annual average concentrations of nitrogen dioxide (NO_2_) and fine particulate matter (PM_2.5_) for 2018 at 1km x 1km grids, modelled to MSOA level using 2011 postcode headcount information^45^.
- Location of care homes: Care homes per 1,000 population using data from the Care Quality Commission via Geolytix^46^.

All covariates were divided into quintiles, giving approximately 1,360 MSOAs in each quintile.

### Statistical methods

All analyses were carried out for males and females separately. We split age into four groups: 40-59 years; 60-69 years; 70-79 years; 80+ years.

We used a two-stage approach in order that the pandemic and comparison periods were treated as independent and distinct. First, we obtained estimates of the death rates in each MSOA for the comparison period of 1 March to 31 May 2015-2019 using a model that incorporated spatial and age terms to obtain stable estimates of death rates in each age group. Then in a second stage, we modelled the death rates from 1 March to 31 May 2020, relative to the death rates estimated for the comparison period. We estimated excess mortality for each MSOA by comparing death rates for these three months between 2020 and 2015-2019 by sex and age-group. In the second stage we included spatial and age terms as well as community variables to assess their effect on excess mortality. The spatial terms in both stages allowed for local smoothing across communities as well as global smoothing across England and were shared across all age groups.

In the first stage we adjusted for age and smoothed over space to obtain stable estimates of the death rates for the comparison period. We assumed that the number of deaths *y*_*itk*_for the i-th MSOA (i=1,…6,791), the t-th year (t=2015,…,2019) and k-th age group (k=40-59, 60-69, 70-79, 80+) arose from a Poisson distribution:

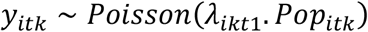

with the log-transformed death rates modelled as a sum of space, age and time terms:

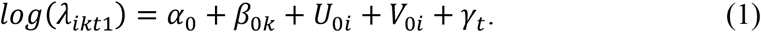

The common intercept for log-transformed death rates is represented by *α*_*0*_, with *β*_*0k*_ the age effect for the*k-th* age group. We modelled MSOA-level intercepts using a Besag, York and Mollie spatial model ^47^; this includes spatially unstructured, independent and identically distributed Gaussian random effects (*V*_0*i*_) and spatially structured random effects (*U*_0*i*_). The latter were modelled with an intrinsic conditional autoregressive prior, which allows for death rates to be more similar across neighbouring MSOAs than those that are far away. This spatial model provides both local and global smoothing on the underlying death rate *λ*_*ikt*1_.

We obtained the posterior distributions of the death rates in each MSOA, age group and year, *λ*_*ikt*1_, and averaged over March to May for the 5 years of the comparison period (2015-2019) to obtain *λ*_*ik*1_, the expected death rate for the *i*-th MSOA and *k*-th age group during March-May 2020 had there been an absence of the pandemic.

In the second stage we estimated the ratio between death rates in March to May 2020 and the death rates we would have expected had there been no pandemic, using data for the same three months for 2015-2019. We estimated the effect of community variables on this ratio. For the number of deaths in the i-th MSOA and k-th age group in 2020, we specified the following model:

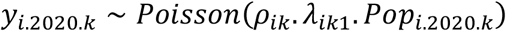

where *ρ*_*ik*_represents the age-specific ratio between death rates in 2020 and the comparison period (*λ*_*ik*1_).

We modelled the ratio *ρ*_*ik*_in a similar way to stage one using terms to account for both space and age:

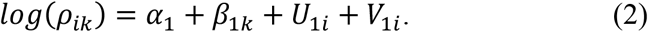

Community variables were incorporated into this second stage log-linear model to evaluate their effect on the mortality rate ratio. For univariable effects we added the term *δX*_*i*_ where *X*_*i*_is the quintile of the variable in the i-th MSOA and *δ* is the associated effect. Similarly for the full multivariable model evaluating the joint effect of all variables we added ∑_*j*_ *δ*_*j*_*X*_*ij*_ withj=1,…6.

To ensure uncertainty in the estimation of *λ*_*ik*1_in stage 1 is expressed in stage 2, we drew 50 samples from the posterior distribution of each *λ*_*ik*1_and ran a stage 2 analysis fixing *λ*_*ik*1_ to each of these values in turn. For each of these 50 analyses we sampled 100 values from the posterior distribution of each *λ*_*ik*2_. In this way we fully expressed the uncertainty resulting from the two stages of our analysis. For the neighbourhood variable effects, we report posterior mean and 95% credible intervals (2.5th to 97.5th percentiles) based on the 5,000 sampled values (50 x 100). In addition, for each MSOA we report both excess deaths per 100,000 people and the percentage change in deaths, as described below.

### Excess deaths per 100,000 people

We obtained the posterior distribution of the estimated number of deaths across ages for March – May 2020, calculated as *ŷ* _*i*.2020_. = ∑ _*k*_*λ*_*ik*1_ × *ρ*_*ik*_× *Pop*_*i*.2020.*k*_, and subtracted the corresponding number for 2015-2019, calculated as *ŷ*_*i*.2015–2019_. = ∑ _*k*_*λ*_*ik*1_ × *Pop*_*i*.2015–2019.*k*_. This difference was then divided by the 2020 population over 40 years old in that MSOA and multiplied by 100,000. Fig. 3(A) shows the excess deaths in map form: the colour key on the maps is categorical such that all MSOAs with excess deaths above 500 per 100,000 are coloured darkest red.

### Percentage change in deaths

We obtained the posterior distribution of the estimated number of deaths across ages for March – May 2020 as summed over the age groups, as *ŷ*_*i*.2020_. = ∑ _*k*_*λ*_*ik*1_ × *ρ*_*ik*_× *Pop*_*i*.2020.*k*_ and divided by the corresponding number for 2015-2019 as *ŷ*_*i*.2015–2019_. = ∑ _*k*_*λ*_*ik*1_ × *Pop*_*i*.2015–2019.*k*_. We then subtracted 1 and multiplied by 100

We fitted the models using the Integrated Nested Laplace Approximation (INLA), through the R-INLA software package (http://www.r-inla.org/)^48,49^. We specified a minimally informative prior, logGamma(1, 0.1), on the hyperparameters τ_V_ and τ_U_.

As sensitivity analyses we re-ran the model using alternative priors for the hyperparameters τ_V_ and τ_U_ firstly using logGamma(0.5, 0.05), and secondly using the penalised complexity prior, as described by Moraga^50^. The results from these sensitivity analyses (Extended Data Table 7a and 7b) show little difference to the main model.

We carried out an analysis in which deaths in care homes were excluded (Extended Data Table 7c and 7d). Finally, we carried out an analysis in which deaths from both sexes were combined (Extended Data Table 7e).

### Ethics and governance

The study was covered by national research ethics approval from the London-South East Research Ethics Committee (Reference 17/LO/0846). Data access was covered by the Health Research Authority Confidentiality Advisory Group under section 251 of the National Health Service Act 2006 and the Health Service (Control of Patient Information) Regulations 2002 (Reference 20/CAG/0008).

### Data availability

- No identifiable information will be shared with any other organisation. SAHSU does not have permission to supply data to third parties. Individual mortality data can be requested through the Office for National Statistics (https://www.ons.gov.uk/).
- Mid-year population estimates can be downloaded from https://www.ons.gov.uk/peoplepopulationandcommunity/populationandmigration/populationestimates/datasets/middlesuperoutputareamidyearpopulationestimates.
- English Index of Multiple Deprivation data can be downloaded from https://www.gov.uk/government/statistics/english-indices-of-deprivation-2019.
- 2011 Census data can be downloaded from https://www.ons.gov.uk/census/2011census/2011censusdata.
- Modelled air pollution data (NO_2_ & PM_2.5_) can be downloaded from https://uk-air.defra.gov.uk/data/pcm-data.
- Locations data of care homes can be downloaded from https://covid19.esriuk.com/datasets/e4ffa672880a4facaab717dea3cdc404_0.

### Code availability

The computer code written in R^51^ for the two stages of Bayesian models used in this work is available on request.

## Acknowledgements

We thank Hima Daby, Gajanan Natu and Eric Johnson for their assistance in data acquisition, storage, preparation and governance; Aubrianna Zhu for her work on the on-line interactive map; and the Office for National Statistics (www.ons.gov.uk) for the provision of mortality data derived from the national mortality registrations.

P.E. is Director of the UK Small Area Health Statistics Unit. He acknowledges support from the Medical Research Council (MRC) for the MRC Centre for Environment and Health (MR/S019669/1); the British Heart Foundation Imperial College Centre for Research Excellence (RE/18/4/34215); National Institute for Health Research (NIHR) Imperial College Biomedical Research Centre (BRC); the NIHR Health Protection Research Unit (HPRU) on Environmental Exposures and Health (NIHR-200880); and the NIHR HPRU in Chemical and Radiation Threats and Hazards (NIHR-200922). P.E. is supported by Health Data Research UK (HDR UK) and the UK Dementia Research Institute at Imperial College London, which receives funding from the UK Medical Research Council, Alzheimer’s Society and Alzheimer’s Research UK (MC_PC_17114). P.E. also acknowledges support from the Huo Family Foundation for research into COVID-19.

J.B. and M.E are supported by Pathways to Equitable Healthy Cities grant from the Wellcome Trust (209376/Z/17/Z).

B.D acknowledges funding from the NIHR HPRU in Chemical and Radiation Threats and Hazards (NIHR-200922) and the HDR UK Hub DISCOVER-NOW.

All authors acknowledge infrastructure support for the Department of Epidemiology and Biostatistics provided by the NIHR Imperial BRC.

The views expressed are those of the authors and not necessarily those of the NIHR or the Department of Health and Social Care.

## Author contributions

P.E and M.E conceived and supervised the study. B.D. and M.B. developed the initial study protocol. M.B. and J.B. developed the statistical model. D.F. prepared the denominator and covariate data. B.D. led the acquisition of identifiable data and study permissions. B.P. authored the computer code, performed the analysis and prepared the initial results. All authors contributed to the drafting of the paper and critical interpretation of results and approve the final version for publication.

## Additional information

Supplementary information is available for this paper.

Correspondence and requests for materials should be addressed to P.E.

## References

1 Kontis, V. et al. Magnitude, demographics and dynamics of the impact of the first phase of the Covid-19 pandemic on all-cause mortality in 17 industrialised countries. Nat Med, doi:https://doi.org/10.1038/s41591-020-1112-0 (2020).

2 Williamson, E. J. et al. Factors associated with COVID-19-related death using OpenSAFELY. Nature 584, 430–436 (2020).

3 Bilinski, A. & Emanuel, E. J. COVID-19 and Excess All-Cause Mortality in the US and 18 Comparison Countries. Jama, doi:10.1001/jama.2020.20717 (2020).

4 Douglas, M., Katikireddi, S. V., Taulbut, M., McKee, M. & McCartney, G. Mitigating the wider health effects of covid-19 pandemic response. 369, m1557, doi:10.1136/bmj.m1557 %J BMJ (2020).

5 Woolf, S. H., Chapman, D. A., Sabo, R. T., Weinberger, D. M. & Hill, L. Excess Deaths From COVID-19 and Other Causes, March-April 2020. Jama 324, 510–513, doi:10.1001/jama.2020.11787 (2020).

6 Office for National Statistics. Analysis of all-cause mortality patterns of selected European countries and regions,week ending 3 January (Week 1) to week ending 12 June (Week 24) 2020., (2020).

7 Zhou, F. et al. Clinical course and risk factors for mortality of adult inpatients with COVID-19 in Wuhan, China: a retrospective cohort study. The Lancet 395, 1054–1062, doi:https://doi.org/10.1016/S0140-6736(20)30566-3 (2020).

8 Propper Carol, S., George and Zaranko, Ben.. The wider impacts of the coronavirus pandemic on the NHS: Briefing note Report No. ISBN 978-1-912805-70-9, (2020).

9 Office for National Statistics. Deaths involving COVID-19 by local area and socioeconomic deprivation: deaths occurring between 1 March and 31 July 2020. (Office for National Statistics, 2020).

10 CDC COVID-19 Response Team. Geographic Differences in COVID-19 Cases, Deaths, and Incidence - United States, February 12-April 7, 2020. MMWR Morb Mortal Wkly Rep. 69, 465–471, doi:10.15585/mmwr.mm6915e4 (2020).

11 Hernández-Moralez, A., Oroschakoff, Kalina & Barigazzi, Jacopo. in Politico (2020).

12 Ward, H. et al. Antibody prevalence for SARS-CoV-2 in England following first peak of the pandemic: REACT2 study in 100,000 adults. medRxiv (2020).

13 Cole, M. A., Ozgen, C. & Strobl, E. Air Pollution Exposure and Covid-19 in Dutch Municipalities. Environmental and Resource Economics 76, 581–610 (2020).

14 Konstantinoudis, G. et al. Long-term exposure to air-pollution and COVID-19 mortality in England: a hierarchical spatial analysis. medRxiv (2020).

15 Magazzino, C., Mele, M. & Schneider, N. The relationship between air pollution and COVID-19-related deaths: an application to three French cities. Applied Energy, 115835 (2020).

16 Wu, X., Nethery, R. C., Sabath, B. M., Braun, D. & Dominici, F. Exposure to air pollution and COVID-19 mortality in the United States. medRxiv (2020).

17 Iacobucci, G. Covid-19: UK government is urged to publish daily care home deaths as it promises more testing. BMJ 369m, 1504, doi:https://doi.org/10.1136/bmj.m1504 (2020).

18 Karanikolos, M. & McKee, M. How comparable is COVID-19 mortality across countries. COVID-19 Health System Monitor 4 (2020).

19 Newton, J. Behind the headlines: counting COVID-19 deaths. Public Health Matters (2020).

20 Watson, J., Whiting, P. F. & Brush, J. E. Interpreting a covid-19 test result. BMJ 369m, 1808, doi:doi: https://doi.org/10.1136/bmj.m1808 (2020).

21 Stevens, S., Pritchard, A.. Important and urgent - next steps on NHS reponse to COVID-19, <https://www.england.nhs.uk/coronavirus/wp-content/uploads/sites/52/2020/03/urgent-next-steps-on-nhs-response-to-covid-19-letter-simon-stevens.pdf> (2020).

22 Emmerson, C. et al. Risk factors for outbreaks of COVID-19 in care homes following hospital discharge: a national cohort analysis. medRxiv (2020).

23 Healthwatch. 590 people’s stories of leaving hospital during COVID-19. (2020).

24 Public Health England. Beyond the data: Understanding the impact of COVID-19 on BAME groups., (PHE, London, UK., 2020).

25 Price-Haywood, E. G., Burton, J., Fort, D. & Seoane, L. Hospitalization and Mortality among Black Patients and White Patients with Covid-19. 382, 2534–2543, doi:10.1056/NEJMsa2011686 (2020).

26 Aldridge, R. W. et al. Black, Asian and Minority Ethnic groups in England are at increased risk of death from COVID-19: indirect standardisation of NHS mortality data. Wellcome Open Research 5, 88 (2020).

27 Patel, J. et al. Poverty, inequality and COVID-19: the forgotten vulnerable. Public health 183, 110 (2020).

28 Smith, L. E. et al. Factors associated with adherence to self-isolation and lockdown measures in the UK; a cross-sectional survey. medRxiv (2020).

29 Stoye, G., Zaranko, B., Shipley, M., McKee, M. & Brunner, E. J. Educational Inequalities in Hospital Use Among Older Adults in England, 2004-2015. The Milbank Quarterly (2020).

30 Mathur, R. et al. Ethnic differences in COVID-19 infection, hospitalisation, and mortality: an OpenSAFELY analysis of 17 million adults in England. medRxiv (2020).

31 House, N., Holborn, H. & Wc, L. ICNARC report on COVID-19 in critical care. Published online 26, 24 (2020).

32 Office for National Statistics. Coronavirus (COVID-19) related deaths by ethnic group, England and Wales: 2 March 2020 to 10 April 2020 [Online].. (2020).

33 Goff, L. M. Ethnicity and Type 2 diabetes in the UK. Diabetic Medicine 36, 927–938 (2019).

34 Townsend, M. J., Kyle, T. K. & Stanford, F. C. Outcomes of COVID-19: disparities in obesity and by ethnicity/race. International Journal of Obesity 44, 1807–1809, doi:https://doi.org/10.1038/s41366-020-0635-2 (2020).

35 Oliver, D. David Oliver: Preventing more deaths in care homes in a second pandemic surge. BMJ 369, m2461, doi:10.1136/bmj.m2461 %J BMJ (2020).

36 Department of Health and Social Care. Local restrictions: areas with an outbreak of coronavirus (COVID-19), <https://www.gov.uk/government/collections/local-restrictions-areas-with-an-outbreak-of-coronavirus-covid-19> (2020).

37 Office for National Statistics. Coronavirus and the impact on output in the UK economy: April 2020. (ONS, 2020).

38 Riley, S., Ainslie, K., Eales, O., Walters, C., Wang, W., Atchison, C., Diggle, P., Ashby, D., Donnelly, C., Cooke, G., Barclay, W., Ward, H., Darzi, D., Elliot, P.. Transient dynamics of SARS-CoV-2 as England exited national lockdown. Preprint (2020).

39 Vinceti, M. et al. Lockdown timing and efficacy in controlling COVID-19 using mobile phone tracking. EClinicalMedicine 25, 100457, doi:10.1016/j.eclinm.2020.100457 (2020).

40 Roderick, P., Macfarlane, A. & Pollock, A. M. Getting back on track: control of covid-19 outbreaks in the community. bmj 369 (2020).

41 Michie, S. et al. Reducing SARS-CoV-2 transmission in the UK: A behavioural science approach to identifying options for increasing adherence to social distancing and shielding vulnerable people. British Journal of Health Psychology (2020).

42 Smith, L. E. et al. Adherence to the test, trace and isolate system: results from a time series of 21 nationally representative surveys in the UK (the COVID-19 Rapid Survey of Adherence to Interventions and Responses [CORSAIR] study). medRxiv (2020).

## Methods References

43 McLennan, D. et al. The English Indices of Deprivation 2019: technical report. (2019).

44 Office for National Statistics. 2011 Census aggregate data. UK Data Service (2016).

45 Department for Environment Food and Rural Affairs. Modelled background pollution data.. (DEFRA., 2020).

46 Geolytix. Geolytix UK Care Homes 2020. (2020).

47 Besag J Y. J., Mollie A. Bayesian image restoration with two applications in spatial statistics. Annals of the Institute of Statistical Mathematics., 1-59 (1991).

48 Blangiardo, M. & Cameletti, M. Spatial and spatio-temporal Bayesian models with R-INLA. (John Wiley & Sons, 2015).

49 Rue, H., Martino, S. & Chopin, N. Approximate Bayesian inference for latent Gaussian models by using integrated nested Laplace approximations. Journal of the royal statistical society: Series b (statistical methodology) 71, 319–392 (2009).

50 Moraga, P. Geospatial Health Data: Modeling and Visualization with R-INLA and Shiny. (Chapman & Hall, 2019).

51 R Core Team. A language and environment for statistical computing [Internet]. Vienna, Austria: R Foundation for Statistical Computing; 2015. Document freely available on the internet at: http://www.r-project.org (2015)

