## Extended data for "Community factors and excess mortality in first wave of the COVID-19 pandemic"

**Extended Data Table 1. Characteristics of the 6,791 Middle Super Output Areas in England.**

| | Mean | Median | Range | Inter-quartile range |
|---|---|---|---|---|
| Population | 8,288 | 7,992 | 2,224 – 24,795 | 6,853 – 9,338 |
| Population (M) | 4,098 | 3,928 | 1,089 – 13,648 | 3,362 – 4,631 |
| Population (F) | 4,191 | 4,058 | 1,135 – 11,147 | 3,476 – 4,714 |
| Population over 40 (M) | 1,994 | 1,930 | 410 – 4,466 | 1,662 – 2,275 |
| Population over 40 (F) | 2,155 | 2,089 | 286 – 4,808 | 1,788 – 2,470 |
| Area ($km^2$) | 19.2 | 3.04 | 0.294 – 1128 | 1.69 – 10.5 |
| Baseline deaths (M), per 100,000 males over 40, 2015 – 2019 | 447 | 443 | 131 – 1,118 | 375 – 513 |
| Baseline deaths (F), per 100,000 females over 40, 2015 – 2019 | 429 | 413 | 118 – 1,419 | 339 – 505 |

M – male; F – female; km- kilometre.

**Extended Data Table 2. Characteristics of the study and comparison populations.**

| | Comparison period (1 March – 31 May, 2015 – 2019) | Study period (1 March – 31 May 2020) | | |
|---|---|---|---|---|
| | **Mean deaths per year (%)** | **Total deaths (%)** | **Deaths with COVID-19 as underlying cause of death (%)** | **Deaths with COVID-19 mentioned on death certificate (%)** |
| **Total** | 121,358 | 171,294 | 41,833 | 44,666 |
| **Sex** | | | | |
| Male | 59,347 (48.9) | 86,071 (50.2) | 23,222 (55.5) | 24,736 (55.4) |
| Female | 62,011 (51.1) | 85,223 (49.8) | 18,611 (44.5) | 19,930 (44.6) |
| **Age** | | | | |
| 40-59 | 10,296 (8.48) | 11,876 (6.93) | 2,505 (5.99) | 2,728 (6.1) |
| 60-69 | 14,287 (11.8) | 17,763 (10.4) | 4,074 (9.74) | 4,377 (9.8) |
| 70-79 | 26,961 (22.2) | 38,378 (22.4) | 9,430 (22.5) | 10,133 (22.7) |
| 80+ | 69,814 (57.5) | 103,277 (60.3) | 25,824 (61.7) | 27,428 (61.4) |
| **Place of death** | | | | |
| Hospital | 56,735 (46.8) | 69,068 (40.3) | 26,755 (64.0) | 28,380 (63.5) |
| Care home | 27,123 (22.3) | 52,112 (30.4) | 12,598 (30.1) | 13,281 (29.7) |
| Home | 28,294 (23.3) | 40,678 (23.7) | 1,776 (4.24) | 2,028 (4.54) |
| Hospice | 6,907 (5.69) | 6,348 (3.71) | 382 (0.91) | 617 (1.38) |
| Other/elsewhere | 2,299 (1.89) | 3,088 (1.80) | 322 (0.77) | 360 (0.81) |
| **Population on income support (%)** | | | | |
| ≥ 0.80 to < 6.00 | 22,877 (18.9) | 32,743 (19.1) | 7,715 (18.4) | 8,251 (18.5) |
| ≥ 6.00 to < 8.79 | 25,047 (20.6) | 34,341 (20.0) | 7,603 (18.2) | 8,146 (18.2) |
| ≥ 8.79 to < 12.7 | 25,272 (20.8) | 35,021 (20.4) | 8,111 (19.4) | 8,690 (19.5) |
| ≥ 12.7 to < 18.8 | 24,435 (20.1) | 34,561 (20.2) | 9,045 (21.6) | 9,611 (21.5) |
| ≥ 18.8 to < 48.9 | 23,727 (19.6) | 34,628 (20.2) | 9,359 (22.4) | 9,968 (22.3) |
| **Population density (population per km$^2$)** | | | | |
| ≥ 5.65 to < 456 | 26,569 (21.9) | 35,011 (20.4) | 6,641 (15.9) | 7,102 (15.9) |
| ≥ 456 to < 1,877 | 27,214 (22.4) | 37,532 (21.9) | 8,527 (20.4) | 9,112 (20.4) |
| ≥ 1,877 to < 3,445 | 25,429 (21.0) | 35,538 (20.7) | 8,619 (20.6) | 9,222 (20.6) |
| ≥ 3,445 to < 5,340 | 24,246 (20.0) | 34,926 (20.4) | 9,034 (21.6) | 9,672 (21.7) |
| ≥ 5,340 to < 29,000 | 17,900 (14.7) | 28,287 (16.5) | 9,015 (21.5) | 9,558 (21.4) |
| **Population non-White (%)** | | | | |
| ≥ 0.43 to < 2.10 | 28,373 (23.4) | 36,559 (21.3) | 6,896 (16.5) | 7,363 (16.5) |
| ≥ 2.10 to < 3.72 | 27,412 (22.6) | 36,974 (21.6) | 7,967 (19.0) | 8,563 (19.2) |
| ≥ 3.72 to < 8.28 | 25,513 (21.0) | 34,834 (20.3) | 8,044 (19.2) | 8,598 (19.2) |
| ≥ 8.28 to < 22.2 | 22,401 (18.5) | 33,010 (19.3) | 8,729 (20.9) | 9,323 (20.9) |
| ≥ 22.2 to < 94.4 | 17,659 (14.6) | 29,917 (17.5) | 10,197 (24.4) | 10,819 (24.2) |
| **Overcrowded homes (%)** | | | | |
| ≥ 0.334 to < 1.65 | 26,425 (21.8) | 35,553 (20.8) | 7,332 (17.5) | 7,869 (17.6) |
| ≥ 1.65 to < 2.42 | 26,769 (22.1) | 36,170 (21.1) | 7,747 (18.5) | 8,314 (18.6) |
| ≥ 2.42 to < 3.67 | 26,564 (21.9) | 36,282 (21.2) | 8,353 (20.0) | 8,923 (20.0) |
| ≥ 3.67 to < 6.26 | 23,979 (19.8) | 34,031 (19.9) | 8,622 (20.6) | 9,209 (20.6) |

| | | | | |
|---|---|---|---|---|
| ≥ 6.26 to < 36.5 | 17,612 (14.5) | 29,258 (17.1) | 9,779 (23.4) | 10,351 (23.2) |
| **$NO_2$ annual average (μg/m$^3$)** | | | | |
| ≥ 3.60 to < 9.52 | 28,640 (23.6) | 36,695 (21.4) | 6,393 (15.3) | 6,868 (15.4) |
| ≥ 9.52 to < 12.3 | 26,833 (22.1) | 36,275 (21.2) | 7,898 (18.9) | 8,441 (18.9) |
| ≥ 12.3 to < 14.9 | 24,449 (20.1) | 33,997 (19.8) | 8,110 (19.4) | 8,701 (19.5) |
| ≥ 14.9 to < 18.7 | 23,884 (19.7) | 34,896 (20.4) | 9,422 (22.5) | 10,051 (22.5) |
| ≥ 18.7 to < 47.5 | 17,553 (14.5) | 29,431 (17.2) | 10,010 (23.9) | 10,605 (23.7) |
| **$PM_{2.5}$ annual average (μg/m$^3$)** | | | | |
| ≥ 5.05 to < 7.76 | 27,660 (22.8) | 36,543 (21.3) | 7,731 (18.5) | 8,286 (18.6) |
| ≥ 7.76 to < 8.91 | 26,173 (21.6) | 35,901 (21.0) | 8,253 (19.7) | 8,796 (19.7) |
| ≥ 8.91 to < 9.88 | 26,131 (21.5) | 35,491 (20.7) | 7,743 (18.5) | 8,338 (18.7) |
| ≥ 9.88 to < 10.9 | 23,709 (19.5) | 33,929 (19.8) | 8,207 (19.6) | 8,760 (19.6) |
| ≥ 10.9 to < 14.4 | 17,684 (14.6) | 29,430 (17.2) | 9,899 (23.7) | 10,486 (23.5) |
| **Care homes per 1,000 population** | | | | |
| 0 | 19,457 (16.0) | 26,536 (15.5) | 6,678 (15.0) | 7,157 (16.0) |
| ≥ 0.040 to < 0.145 | 19,431 (16.0) | 27,553 (16.2) | 6,993 (16.7) | 7,502 (16.8) |
| ≥ 0.145 to < 0.265 | 24,163 (19.7) | 34,577 (20.2) | 8,732 (20.9) | 9,282 (20.8) |
| ≥ 0.265 to < 0.438 | 27,229 (22.4) | 38,375 (22.4) | 9,161 (21.9) | 9,736 (21.8) |
| ≥ 0.438 to < 4.23 | 31,078 (25.6) | 43,253 (25.8) | 10,269 (24.5) | 10,989 (24.6) |

**Extended Data Table 3. Excess death rates, percent increase in mortality and posterior probabilities for Middle Super Output Areas in England.**

| | Mean | Median | Range | Inter-quartile range |
|---|---|---|---|---|
| Excess death rate (M), per 100,000 males over 40 | 206 | 182 | -248 – 1488 | 96.2 – 288 |
| Excess death rate (F), per 100,000 females over 40 | 166 | 132 | -272 - 2550 | 55.2 - 237 |
| Percent increase in mortality (M) | 51.2 | 45.0 | -36.9 – 288 | 24.7 – 70.2 |
| Percent increase in mortality (F) | 42.2 | 36.3 | -41.8 - 302 | 16.4 – 61.6 |
| Posterior probability that excess rate is greater than zero (M) | 0.872 | 0.948 | 0.032 – 1.00 | 0.819 – 0.992 |
| Posterior probability that excess rate is greater than zero (F) | 0.803 | 0.882 | 0.009 – 1.00 | 0.969 – 0.973 |

M – male; F – female.

**Extended Data Table 4. One-variable-at-a-time models for community characteristics.** Age is included in all the models.

| Variable | n-tile | Males | | Females | |
|---|---|---|---|---|---|
| | | Mean | 95% credible interval | Mean | 95% credible interval |
| **Population on income support** | 1 (lowest) | ref | ref | ref | ref |
| | 2 | 1.05 | 1.01 – 1.08 | 0.99 | 0.96 – 1.03 |
| | 3 | 1.10 | 1.06 – 1.13 | 1.03 | 1.00 – 1.07 |
| | 4 | 1.15 | 1.11 – 1.18 | 1.06 | 1.02 – 1.09 |
| | 5 (highest) | 1.21 | 1.17 – 1.25 | 1.15 | 1.11 – 1.19 |
| | % of variation contributed by variable | 5.66 | 4.15 – 7.35 | 3.28 | 2.17 – 4.65 |
| **Population density** | 1 (lowest) | ref | ref | ref | ref |
| | 2 | 1.06 | 1.03 – 1.09 | 1.04 | 1.01 – 1.07 |
| | 3 | 1.09 | 1.06 – 1.13 | 1.04 | 1.01 – 1.08 |
| | 4 | 1.10 | 1.06 – 1.13 | 1.09 | 1.05 – 1.13 |
| | 5 (highest) | 1.16 | 1.11 – 1.20 | 1.11 | 1.06 – 1.15 |
| | % of variation contributed by variable | 3.20 | 1.77 – 4.90 | 1.71 | 0.784 – 2.88 |
| **Population non-White** | 1 (lowest) | ref | ref | ref | ref |
| | 2 | 1.04 | 1.01 – 1.07 | 1.04 | 1.01 – 1.08 |
| | 3 | 1.06 | 1.03 – 1.10 | 1.04 | 1.01 – 1.08 |
| | 4 | 1.12 | 1.08 – 1.16 | 1.12 | 1.08 – 1.16 |
| | 5 (highest) | 1.26 | 1.20 – 1.31 | 1.21 | 1.15 – 1.26 |
| | % of variation contributed by variable | 8.80 | 5.93 – 11.9 | 5.32 | 3.15 – 7.73 |
| **Overcrowded homes** | 1 (lowest) | ref | ref | ref | ref |
| | 2 | 1.04 | 1.01 – 1.07 | 1.02 | 0.99 – 1.05 |
| | 3 | 1.08 | 1.05 – 1.12 | 1.05 | 1.02 – 1.09 |
| | 4 | 1.15 | 1.11 – 1.18 | 1.09 | 1.06 – 1.13 |
| | 5 (highest) | 1.26 | 1.22 – 1.31 | 1.18 | 1.14 – 1.23 |
| | % of variation contributed by variable | 9.21 | 6.64 – 12.1 | 4.44 | 2.68 – 6.50 |
| **$NO_2$** | 1 (lowest) | ref | ref | ref | ref |
| | 2 | 1.04 | 1.01 – 1.08 | 1.04 | 1.00 – 1.07 |
| | 3 | 1.09 | 1.05 – 1.13 | 1.04 | 1.01 – 1.08 |
| | 4 | 1.13 | 1.09 – 1.17 | 1.11 | 1.07 – 1.16 |
| | 5 (highest) | 1.22 | 1.16 – 1.28 | 1.17 | 1.11 – 1.23 |
| | % of variation contributed by variable | 6.42 | 3.75 – 9.64 | 3.92 | 1.98 – 6.30 |
| **$PM_{2.5}$** | 1 (lowest) | ref | ref | ref | ref |
| | 2 | 1.06 | 1.02 – 1.10 | 1.04 | 1.00 – 1.08 |
| | 3 | 1.12 | 1.07 – 1.18 | 1.06 | 1.01 – 1.12 |
| | 4 | 1.17 | 1.10 – 1.23 | 1.10 | 1.04 – 1.16 |
| | 5 (highest) | 1.28 | 1.20 – 1.37 | 1.19 | 1.10 – 1.27 |
| | % of variation contributed by variable | 9.30 | 5.27 – 14.13 | 4.12 | 1.50 – 7.55 |

| | | | | | |
|---|---|---|---|---|---|
| **Care home per 1,000 population** | 1 (lowest) | ref | ref | ref | ref |
| | 2 | 1.10 | 1.07 – 1.13 | 1.13 | 1.09 – 1.17 |
| | 3 | 1.13 | 1.09 – 1.16 | 1.18 | 1.14 – 1.22 |
| | 4 | 1.15 | 1.12 – 1.18 | 1.21 | 1.18 – 1.25 |
| | 5 (highest) | 1.21 | 1.17 – 1.25 | 1.27 | 1.23 – 1.31 |
| | % of variation contributed by variable | 4.61 | 3.43 – 5.81 | 6.66 | 5.17 – 8.14 |

$NO_2$ – nitrogen dioxide; PM2.5 – particulate matter 2.5 μ diameter

**Extended Data Table 5. Correlation between community characteristics for Middle Super Output Areas in England:** Kendall's Tau coefficients between covariates (quintiles)

| | Population on income support | Population density | Non-White population | Overcrowding | $NO_2$ | $PM_{2.5}$ | Care homes per 1000 population* |
|---|---|---|---|---|---|---|---|
| **Population on income support** | 1 | | | | | | |
| **Population density** | 0.37 | 1 | | | | | |
| **Population non-White** | 0.21 | 0.56 | 1 | | | | |
| **Overcrowding** | 0.55 | 0.61 | 0.61 | 1 | | | |
| **$NO_2$** | 0.26 | 0.59 | 0.67 | 0.56 | 1 | | |
| **$PM_{2.5}$** | 0.04 | 0.39 | 0.55 | 0.42 | 0.55 | 1 | |
| **Care homes per 1000 population*** | -0.04 | -0.17 | -0.17 | -0.16 | -0.20 | -0.16 | 1 |

*All MSOAs with a care home density of 0 (n=1,535) are in quintile 1. Quintile 2 has reduced number of MSOAs (n=1,182)

**Extended Data Table 6. Mutually adjusted multivariable models for all MSOA characteristics.** Age is included in all the models.

| **Variable** | **Quintile** | **Males** | | **Females** | |
|---|---|---|---|---|---|
| | | **Mean** | **95% credible interval** | **Mean** | **95% credible interval** |
| **Population on income support** | 1 (lowest) | ref | ref | ref | ref |
| | 2 | 1.02 | 0.99 – 1.05 | 0.97 | 0.94 – 1.01 |
| | 3 | 1.04 | 1.01 – 1.08 | 0.99 | 0.95 – 1.03 |
| | 4 | 1.07 | 1.03 – 1.12 | 1.01 | 0.96 – 1.05 |
| | 5 (highest) | 1.10 | 1.05 – 1.16 | 1.08 | 1.02 – 1.13 |
| **Population density** | 1 (lowest) | ref | ref | ref | ref |
| | 2 | 1.02 | 0.97 – 1.04 | 1.01 | 0.97 – 1.04 |
| | 3 | 1.01 | 0.98 – 1.05 | 0.99 | 0.95 – 1.03 |
| | 4 | 0.99 | 0.95 – 1.04 | 1.02 | 0.97 – 1.06 |
| | 5 (highest) | 0.99 | 0.95 – 1.04 | 1.00 | 0.94 – 1.05 |
| **Population non-White** | 1 (lowest) | ref | ref | ref | ref |
| | 2 | 1.01 | 0.98 – 1.04 | 1.03 | 0.99 – 1.06 |
| | 3 | 1.02 | 0.98 – 1.06 | 1.02 | 0.98 – 1.06 |
| | 4 | 1.04 | 0.99 – 1.08 | 1.06 | 1.01 – 1.12 |
| | 5 (highest) | 1.08 | 1.02 – 1.15 | 1.09 | 1.02 – 1.17 |
| **Overcrowded homes** | 1 (lowest) | ref | ref | ref | ref |
| | 2 | 1.01 | 0.98 – 1.05 | 1.01 | 0.98 – 1.04 |
| | 3 | 1.02 | 0.99 – 1.06 | 1.01 | 0.97 – 1.06 |
| | 4 | 1.06 | 1.01 – 1.11 | 1.02 | 0.97 – 1.07 |
| | 5 (highest) | 1.12 | 1.04 – 1.19 | 1.06 | 0.99 – 1.14 |
| **$NO_2$** | 1 (lowest) | ref | ref | ref | ref |
| | 2 | 1.01 | 0.97 – 1.04 | 1.02 | 0.98 – 1.06 |
| | 3 | 1.02 | 0.98 – 1.07 | 1.01 | 0.96 – 1.06 |
| | 4 | 1.01 | 0.95 – 1.06 | 1.04 | 0.98 – 1.10 |
| | 5 (highest) | 1.02 | 0.95 – 1.10 | 1.05 | 0.98 – 1.13 |
| **$PM_{2.5}$** | 1 (lowest) | ref | ref | ref | ref |
| | 2 | 1.00 | 0.96 – 1.05 | 0.99 | 0.95 – 1.04 |
| | 3 | 1.03 | 0.97 – 1.09 | 0.98 | 0.92 – 1.04 |
| | 4 | 1.02 | 0.95 – 1.09 | 0.98 | 0.91 – 1.05 |
| | 5 (highest) | 1.07 | 0.98 – 1.17 | 1.02 | 0.93 – 1.11 |
| **Care homes per 1,000 population** | 1 (lowest | ref | ref | ref | ref |
| | 2 | 1.09 | 1.06 – 1.12 | 1.12 | 1.09 – 1.16 |
| | 3 | 1.13 | 1.10 – 1.17 | 1.19 | 1.15 – 1.22 |
| | 4 | 1.15 | 1.12 – 1.19 | 1.22 | 1.19 – 1.26 |
| | 5 (highest) | 1.22 | 1.19 – 1.26 | 1.27 | 1.23 – 1.31 |
| % of variation contributed by all variables | | 17.6 | 13.9 – 21.9 | 14.9 | 12.1 – 18.2 |
| % of variance explained by local clustering for model with all variables | | 30.8 | 28.6 – 32.9 | 18.4 | 17.2 – 20.1 |
| % of variance explained by local clustering for model with no variables | | 39.0 | 38.6 – 41.1 | 21.9 | 20.8 – 23.1 |

**Extended Data Table 7. Mutually adjusted multivariable models for all MSOA characteristics, results of sensitivity analyses.** Age is included in all the models.

**a) Penalised complexity prior.**

| Variable | Quintile | Males | | Females | |
|---|---|---|---|---|---|
| | | Mean | 95% credible interval | Mean | 95% credible interval |
| **Population on income support** | 1 (lowest) | ref | ref | ref | ref |
| | 2 | 1.02 | 0.99 – 1.05 | 0.97 | 0.94 – 1.01 |
| | 3 | 1.04 | 1.00 – 1.08 | 0.99 | 0.95 – 1.03 |
| | 4 | 1.07 | 1.02 – 1.11 | 1.00 | 0.96 – 1.05 |
| | 5 (highest) | 1.10 | 1.05 – 1.15 | 1.07 | 1.02 – 1.13 |
| **Population density** | 1 (lowest) | ref | ref | ref | ref |
| | 2 | 1.02 | 0.97 – 1.04 | 1.01 | 0.97 – 1.04 |
| | 3 | 1.01 | 0.98 – 1.05 | 0.99 | 0.95 – 1.03 |
| | 4 | 0.99 | 0.95 – 1.04 | 1.02 | 0.97 – 1.06 |
| | 5 (highest) | 0.99 | 0.95 – 1.04 | 0.99 | 0.94 – 1.05 |
| **Population non-White** | 1 (lowest) | ref | ref | ref | ref |
| | 2 | 1.01 | 0.98 – 1.05 | 1.03 | 1.00 – 1.06 |
| | 3 | 1.02 | 0.99 – 1.06 | 1.02 | 0.98 – 1.06 |
| | 4 | 1.04 | 0.99 – 1.09 | 1.07 | 1.01 – 1.12 |
| | 5 (highest) | 1.09 | 1.02 – 1.15 | 1.10 | 1.03 – 1.17 |
| **Overcrowded homes** | 1 (lowest) | ref | ref | ref | ref |
| | 2 | 1.01 | 0.98 – 1.05 | 1.01 | 0.98 – 1.04 |
| | 3 | 1.02 | 0.99 – 1.06 | 1.02 | 0.97 – 1.06 |
| | 4 | 1.06 | 1.01 – 1.11 | 1.02 | 0.97 – 1.08 |
| | 5 (highest) | 1.12 | 1.04 – 1.19 | 1.06 | 0.99 – 1.14 |
| **$NO_2$** | 1 (lowest) | ref | ref | ref | ref |
| | 2 | 1.01 | 0.97 – 1.04 | 1.02 | 0.98 – 1.06 |
| | 3 | 1.02 | 0.98 – 1.07 | 1.01 | 0.96 – 1.06 |
| | 4 | 1.01 | 0.95 – 1.06 | 1.04 | 0.98 – 1.10 |
| | 5 (highest) | 1.02 | 0.95 – 1.10 | 1.06 | 0.98 – 1.13 |
| **$PM_{2.5}$** | 1 (lowest) | ref | ref | ref | ref |
| | 2 | 1.00 | 0.96 – 1.05 | 1.00 | 0.95 – 1.04 |
| | 3 | 1.03 | 0.97 – 1.09 | 0.98 | 0.93 – 1.04 |
| | 4 | 1.02 | 0.95 – 1.10 | 0.99 | 0.92 – 1.06 |
| | 5 (highest) | 1.07 | 0.98 – 1.17 | 1.03 | 0.93 – 1.12 |
| **Care homes per 1,000 population** | 1 (lowest | ref | ref | ref | ref |
| | 2 | 1.09 | 1.06 – 1.12 | 1.12 | 1.09 – 1.16 |
| | 3 | 1.13 | 1.10 – 1.17 | 1.19 | 1.15 – 1.22 |
| | 4 | 1.15 | 1.12 – 1.19 | 1.22 | 1.18 – 1.26 |
| | 5 (highest) | 1.22 | 1.19 – 1.26 | 1.27 | 1.23 – 1.31 |
| % of variation contributed by all variables | | 17.9 | 14.1 – 22.3 | 15.3 | 12.4 – 18.5 |

**b) Priors on the hyperparameters $\tau_v$ and $\tau_u$: logGamma(0.5, 0.05).**

| Variable | Quintile | Males | | Females | |
|---|---|---|---|---|---|
| | | Mean | 95% credible interval | Mean | 95% credible interval |
| **Population on income support** | 1 (lowest) | ref | ref | ref | ref |
| | 2 | 1.02 | 0.99 – 1.05 | 0.97 | 0.94 – 1.00 |
| | 3 | 1.04 | 1.01 – 1.08 | 0.99 | 0.95 – 1.03 |
| | 4 | 1.07 | 1.03 – 1.11 | 1.01 | 0.96 – 1.05 |
| | 5 (highest) | 1.10 | 1.05 – 1.16 | 1.08 | 1.02 – 1.13 |
| **Population density** | 1 (lowest) | ref | ref | ref | ref |
| | 2 | 1.02 | 0.99 – 1.06 | 1.01 | 0.97 – 1.04 |
| | 3 | 1.01 | 0.98 – 1.05 | 0.99 | 0.95 – 1.03 |
| | 4 | 0.99 | 0.95 – 1.03 | 1.02 | 0.97 – 1.06 |
| | 5 (highest) | 1.00 | 0.95 – 1.05 | 0.99 | 0.94 – 1.04 |
| **Population non-White** | 1 (lowest) | ref | ref | ref | ref |
| | 2 | 1.01 | 0.98 – 1.04 | 1.03 | 1.00 – 1.06 |
| | 3 | 1.02 | 0.98 – 1.06 | 1.02 | 0.98 – 1.06 |
| | 4 | 1.03 | 0.99 – 1.08 | 1.07 | 1.02 – 1.12 |
| | 5 (highest) | 1.08 | 1.02 – 1.15 | 1.10 | 1.03 – 1.17 |
| **Overcrowded homes** | 1 (lowest) | ref | ref | ref | ref |
| | 2 | 1.01 | 0.98 – 1.05 | 1.01 | 0.98 – 1.04 |
| | 3 | 1.02 | 0.98 – 1.06 | 1.01 | 0.97 – 1.05 |
| | 4 | 1.06 | 1.01 – 1.11 | 1.02 | 0.97 – 1.08 |
| | 5 (highest) | 1.11 | 1.04 – 1.19 | 1.06 | 0.99 – 1.13 |
| **$NO_2$** | 1 (lowest) | ref | ref | ref | ref |
| | 2 | 1.01 | 0.97 – 1.05 | 1.02 | 0.98 – 1.06 |
| | 3 | 1.02 | 0.98 – 1.07 | 1.01 | 0.96 – 1.06 |
| | 4 | 1.01 | 0.95 – 1.07 | 1.04 | 0.98 – 1.10 |
| | 5 (highest) | 1.03 | 0.96 – 1.10 | 1.05 | 0.98 – 1.13 |
| **$PM_{2.5}$** | 1 (lowest) | ref | ref | ref | ref |
| | 2 | 1.00 | 0.96 – 1.05 | 1.00 | 0.95 – 1.04 |
| | 3 | 1.03 | 0.97 – 1.09 | 0.98 | 0.92 – 1.04 |
| | 4 | 1.02 | 0.95 – 1.09 | 0.98 | 0.91 – 1.06 |
| | 5 (highest) | 1.07 | 0.98 – 1.17 | 1.02 | 0.93 – 1.12 |
| **Care homes per 1,000 population** | 1 (lowest | ref | ref | ref | ref |
| | 2 | 1.09 | 1.06 – 1.12 | 1.12 | 1.09 – 1.16 |
| | 3 | 1.13 | 1.10 – 1.17 | 1.19 | 1.15 – 1.23 |
| | 4 | 1.15 | 1.12 – 1.19 | 1.22 | 1.18 – 1.27 |
| | 5 (highest) | 1.22 | 1.19 – 1.26 | 1.27 | 1.23 – 1.32 |
| % of variation contributed by all variables | | 17.6 | 13.9 – 22.0 | 15.0 | 12.0 – 18.1 |

**c) Excluding care homes per 1,000 population.**

| Variables | Quintile | Males | | Females | |
|---|---|---|---|---|---|
| | | **Mean** | **95% credible interval** | **Mean** | **95% credible interval** |
| **Population on income support** | 1 (lowest) | ref | ref | ref | ref |
| | 2 | 1.03 | 1.00 – 1.06 | 0.98 | 0.95 – 1.02 |
| | 3 | 1.06 | 1.02 – 1.10 | 1.01 | 0.97 – 1.05 |
| | 4 | 1.09 | 1.04 – 1.13 | 1.02 | 0.98 – 1.07 |
| | 5 (highest) | 1.12 | 1.07 – 1.18 | 1.10 | 1.04 – 1.16 |
| **Population density** | 1 (lowest) | ref | ref | ref | ref |
| | 2 | 1.02 | 0.99 – 1.05 | 1.01 | 0.97 – 1.05 |
| | 3 | 1.01 | 0.97 – 1.05 | 0.99 | 0.95 – 1.03 |
| | 4 | 0.98 | 0.94 – 1.02 | 1.01 | 0.96 – 1.05 |
| | 5 (highest) | 0.99 | 0.94 – 1.03 | 0.98 | 0.93 – 1.03 |
| **Population non-White** | 1 (lowest) | ref | ref | ref | ref |
| | 2 | 1.02 | 0.99 – 1.05 | 1.04 | 1.00 – 1.07 |
| | 3 | 1.03 | 0.99 – 1.07 | 1.03 | 0.99 – 1.07 |
| | 4 | 1.05 | 1.00 – 1.10 | 1.08 | 1.03 – 1.13 |
| | 5 (highest) | 1.11 | 1.04 – 1.18 | 1.12 | 1.05 – 1.19 |
| **Overcrowded homes** | 1 (lowest) | ref | ref | ref | ref |
| | 2 | 1.01 | 0.98 – 1.05 | 1.01 | 0.97 – 1.04 |
| | 3 | 1.02 | 0.98 – 1.06 | 1.01 | 0.97 – 1.05 |
| | 4 | 1.05 | 1.00 – 1.10 | 1.01 | 0.96 – 1.06 |
| | 5 (highest) | 1.09 | 1.02 – 1.16 | 1.03 | 0.96 – 1.11 |
| **$NO_2$** | 1 (lowest) | ref | ref | ref | ref |
| | 2 | 1.01 | 0.97 – 1.04 | 1.02 | 0.98 – 1.06 |
| | 3 | 1.01 | 0.97 – 1.06 | 1.00 | 0.95 – 1.05 |
| | 4 | 1.01 | 0.95 – 1.07 | 1.04 | 0.98 – 1.10 |
| | 5 (highest) | 1.02 | 0.95 – 1.10 | 1.04 | 0.97 – 1.12 |
| **$PM_{2.5}$** | 1 (lowest) | ref | ref | ref | ref |
| | 2 | 1.00 | 0.96 – 1.04 | 0.99 | 0.95 – 1.04 |
| | 3 | 1.02 | 0.96 – 1.07 | 0.97 | 0.91 – 1.02 |
| | 4 | 1.00 | 0.93 – 1.07 | 0.96 | 0.91 – 1.02 |
| | 5 (highest) | 1.04 | 0.95 – 1.13 | 0.98 | 0.90 – 1.08 |
| % of variation contributed by all variables | | 12.7 | 9.12 – 16.9 | 8.00 | 5.53 – 11.0 |

**d) Excluding care homes per 1,000 population and deaths in care homes.**

| Covariate | Quintile | Males | | Females | |
|---|---|---|---|---|---|
| | | **Mean** | **95% credible interval** | **Mean** | **95% credible interval** |
| **Population on income support** | 1 (lowest) | ref | ref | ref | ref |
| | 2 | 1.05 | 1.02 – 1.09 | 1.02 | 0.99 – 1.06 |
| | 3 | 1.09 | 1.05 – 1.13 | 1.08 | 1.04 – 1.12 |
| | 4 | 1.12 | 1.07 – 1.17 | 1.13 | 1.08 – 1.18 |
| | 5 (highest) | 1.19 | 1.13 – 1.25 | 1.24 | 1.17 – 1.30 |
| **Population density** | 1 (lowest) | ref | ref | ref | ref |
| | 2 | 1.01 | 0.98 – 1.05 | 1.02 | 0.99 – 1.06 |
| | 3 | 1.01 | 0.97 – 1.05 | 1.01 | 0.97 – 1.05 |
| | 4 | 0.97 | 0.93 – 1.01 | 1.02 | 0.98 – 1.07 |
| | 5 (highest) | 0.97 | 0.92 – 1.02 | 0.99 | 0.94 – 1.04 |
| **Population non-White** | 1 (lowest) | ref | ref | ref | ref |
| | 2 | 1.00 | 0.97 – 1.03 | 1.01 | 0.98 – 1.05 |
| | 3 | 0.99 | 0.95 – 1.03 | 0.97 | 0.94 – 1.02 |
| | 4 | 0.99 | 0.94 – 1.04 | 1.03 | 0.98 – 1.09 |
| | 5 (highest) | 1.06 | 1.00 – 1.13 | 1.05 | 0.98 – 1.12 |
| **Overcrowded homes** | 1 (lowest) | ref | ref | ref | ref |
| | 2 | 1.01 | 0.98 – 1.05 | 1.02 | 0.98 – 1.06 |
| | 3 | 1.03 | 0.98 – 1.07 | 1.00 | 0.96 – 1.04 |
| | 4 | 1.06 | 1.01 – 1.11 | 1.02 | 0.97 – 1.07 |
| | 5 (highest) | 1.10 | 1.03 – 1.18 | 1.07 | 1.00 – 1.15 |
| **$NO_2$** | 1 (lowest) | ref | ref | ref | ref |
| | 2 | 1.01 | 0.97 – 1.05 | 0.99 | 0.95 – 1.03 |
| | 3 | 1.02 | 0.97 – 1.07 | 0.98 | 0.93 – 1.03 |
| | 4 | 1.01 | 0.96 – 1.07 | 1.00 | 0.94 – 1.07 |
| | 5 (highest) | 1.05 | 0.98 – 1.13 | 1.03 | 0.95 – 1.11 |
| **$PM_{2.5}$** | 1 (lowest) | ref | ref | ref | ref |
| | 2 | 1.00 | 0.96 – 1.05 | 0.99 | 0.94 – 1.03 |
| | 3 | 1.02 | 0.96 – 1.08 | 0.96 | 0.90 – 1.02 |
| | 4 | 1.02 | 0.95 – 1.10 | 0.96 | 0.89 – 1.04 |
| | 5 (highest) | 1.05 | 0.96 – 1.16 | 0.98 | 0.89 – 1.08 |
| % of variation contributed by all variables | | 20.0 | 15.1 – 25.7 | 24.0 | 18.5 – 30.0 |

**e) Both sexes combined.**

| Variables | Quintile | Mean | 95% credible interval |
|---|---|---|---|
| **Population on income support** | 1 (lowest) | ref | ref |
| | 2 | 0.99 | 0.97 – 1.02 |
| | 3 | 1.01 | 0.98 – 1.04 |
| | 4 | 1.02 | 0.99 – 1.06 |
| | 5 (highest) | 1.06 | 1.02 – 1.10 |
| **Population density** | 1 (lowest) | ref | ref |
| | 2 | 1.01 | 0.99 – 1.04 |
| | 3 | 1.00 | 0.97 – 1.03 |
| | 4 | 1.01 | 0.97 – 1.04 |
| | 5 (highest) | 1.00 | 0.96 – 1.04 |
| **Population non-White** | 1 (lowest) | ref | ref |
| | 2 | 1.02 | 0.99 – 1.04 |
| | 3 | 1.02 | 0.98 – 1.05 |
| | 4 | 1.05 | 1.01 – 1.09 |
| | 5 (highest) | 1.09 | 1.04 – 1.15 |
| **Overcrowded homes** | 1 (lowest) | ref | ref |
| | 2 | 1.01 | 0.98 – 1.03 |
| | 3 | 1.00 | 0.97 – 1.04 |
| | 4 | 1.02 | 0.98 – 1.06 |
| | 5 (highest) | 1.05 | 0.99 – 1.11 |
| **$NO_2$** | 1 (lowest) | ref | ref |
| | 2 | 1.01 | 0.98 – 1.04 |
| | 3 | 1.01 | 0.98 – 1.05 |
| | 4 | 1.02 | 0.97 – 1.07 |
| | 5 (highest) | 1.04 | 0.98 – 1.10 |
| **$PM_{2.5}$** | 1 (lowest) | ref | ref |
| | 2 | 1.00 | 0.96 – 1.03 |
| | 3 | 1.00 | 0.95 – 1.05 |
| | 4 | 0.99 | 0.94 – 1.06 |
| | 5 (highest) | 1.04 | 0.96 – 1.11 |
| **Care homes per 1,000 population** | 1 (lowest | ref | ref |
| | 2 | 1.09 | 1.07 – 1.12 |
| | 3 | 1.14 | 1.11 – 1.16 |
| | 4 | 1.16 | 1.13 – 1.19 |
| | 5 (highest) | 1.21 | 1.18 – 1.24 |
| % of variation contributed by all variables | | 13.7 | 11.0 – 16.7 |

**Extended Data Fig. 1. Scatter plot showing male vs female percent increase in deaths age 40 and over for 6,701 Middle Super Output Areas in England.** Spearman's rank correlation coefficient 0.43

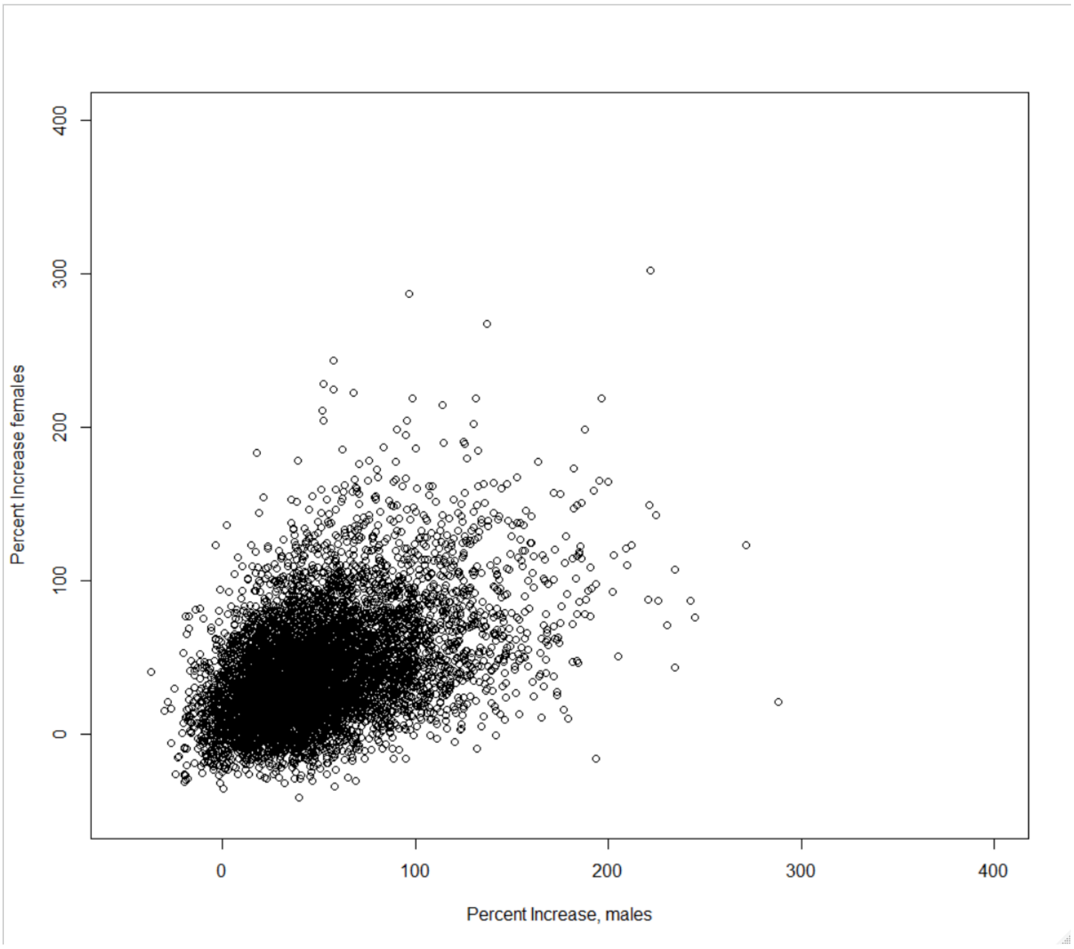

**Extended Data Fig. 2. Scatter plot showing male vs female excess mortality rate per 100,000 males/females age 40 and over for 6,701 Middle Super Output Areas in England.** Spearman's rank correlation coefficient 0.45

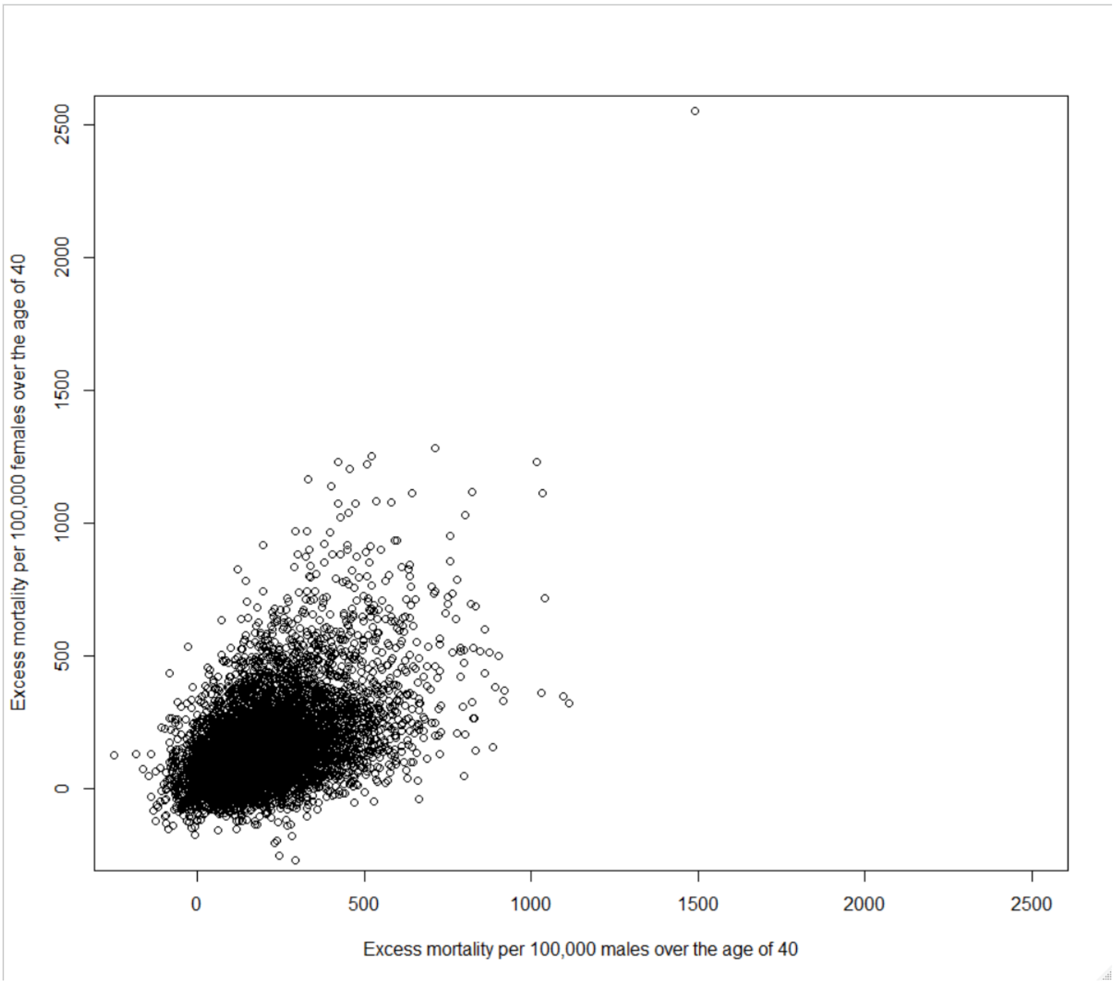
